# Endothelial-protective effects of a G-protein-biased sphingosine-1 phosphate receptor-1 agonist, SAR247799, in type-2 diabetes rats and a randomized placebo-controlled patient trial

**DOI:** 10.1101/2020.05.15.20103101

**Authors:** Luc Bergougnan, Grit Andersen, Leona Plum-Mörschel, Maria Francesca Evaristi, Bruno Poirier, Agnes Tardat, Marcel Ermer, Theresa Herbrand, Jorge Arrubla, Hans Veit Coester, Roberto Sansone, Christian Heiss, Olivier Vitse, Fabrice Hurbin, Rania Boiron, Xavier Benain, David Radzik, Philip Janiak, Anthony J Muslin, Lionel Hovsepian, Stephane Kirkesseli, Paul Deutsch, Ashfaq A Parkar

**Author notes:** **Correspondence**: Ashfaq A Parkar, Global Project Head, Cardiovascular and Diabetes, 55 Corporate Drive, Bridgewater, NJ 08807 USA. The authors confirm that the Principal Investigators for the clinical study were Grit Anderson and Leona Plum-Mörschel and that they had direct clinical responsibility for patients at the Neuss and Mainz sites, respectively. In addition, Grit Andersen was coordinating principal investigator.

## Abstract

SAR247799 is a G-protein-biased sphingosine-1 phosphate receptor-1 (S1P_1_) agonist designed to activate endothelial S1P_1_ and provide endothelial-protective properties, while limiting S1P_1_ desensitization and consequent lymphocyte-count reduction associated with higher doses. A dose-response study in diabetic rats with 5-week SAR247799 treatment demonstrated, at sub-lymphocyte-reducing doses, renal function and endothelial biomarker improvements and was used to select doses for human investigation. Type-2 diabetes patients, enriched for endothelial dysfunction (flow-mediated dilation, FMD<7%) (n=54), were randomized, in two sequential cohorts, to 28-day once-daily treatment with SAR247799 (1 or 5 mg in ascending cohorts), placebo or 50 mg sildenafil (positive control) in a 5:2:2 ratio per cohort. The maximum FMD change from baseline versus placebo for all treatments was reached on day 35; mean differences versus placebo were 0.60% (95% CI –0.34%-1.53%; p=0.203) for 1 mg SAR247799, 1.07% (95% CI 0.13%-2.01%; p=0.026) for 5 mg SAR247799 and 0.88% (95% CI –0.15%-1.91%; p=0.093) for 50 mg sildenafil. Both doses of SAR247799 were well tolerated, did not affect blood pressure, and were associated with minimal-to-no lymphocyte reduction and small-to-moderate heart rate decrease. These data provide the first human evidence for endothelial-protective properties of S1P_1_ activation, with SAR247799 being at least as effective as the clinical benchmark, sildenafil.

**Trial Registration:** www.clinicaltrials.gov (Unique Identifier: NCT03462017; Registered 12/03/2018)

## INTRODUCTION

Endothelial dysfunction is believed to contribute to the progression of a variety of vascular diseases.^1^ It manifests as impaired reactivity of the micro- and/or macro-vasculature to pharmacological or physiological stimuli, as disruption of barrier integrity causing vascular leakage, and as changes in the levels of plasma and/or urinary biomarkers.^2^ These changes are considered to play a major role in the development of atherosclerosis, acute coronary syndromes, stroke and renal damage, and are frequent complications in diabetes.^3^ Diabetes is a pre-disposing factor for worsening cardiovascular outcomes, and indices of endothelial dysfunction can predict such risk.^4^

Sphingosine-1 phosphate (S1P) is a sphingolipid mediator present at high levels in the plasma of healthy individuals, where it is believed to play an important role in maintaining vascular health.^5^ Endothelium and erythrocytes contribute, primarily, to the plasma pool of S1P, and the apoM-component of high-density lipoprotein (HDL) is the major carrier in plasma.^5,6^ Not surprisingly, S1P and HDL-bound S1P are reduced in pathologies or conditions associated with endothelial injury, including diabetes,^7–13^ metabolic syndrome,^14^ myocardial infarction,^12,13^ sepsis,^15,16^ in-stent restenosis,^17^ and in coronary and peripheral artery disease.^18^ In patients with cardiovascular disease, higher S1P is associated with cardio-protective settings such as pre-infarction angina^19^ and coronary collateral circulation.^20^

Although a large body of genetic and epidemiological evidence implicates HDL as a cardio-protective agent, multiple outcome trials with HDL-elevating drugs have provided disappointing results.^21–23^ It has been proposed that this apparent discrepancy can be explained by the quality (or functional capacity) of HDL particles being the critical factor affecting cardiovascular outcomes rather than the absolute HDL level.^21,24^ In this regard, the endothelial-protective properties of HDL are mediated by its S1P cargo, acting via S1P receptor-1 (S1P_1_).^25–29^ Dysfunctional HDL from atherosclerosis and/or diabetes patients has reduced S1P,^18^ and the endothelial barrier-promoting properties of dysfunctional HDL can be restored by S1P_1_ activation.^30^ In preclinical models, S1P_1_ activation maintains endothelial function whereas S1P_1_ inactivation causes endothelial damage.^31,32^ Lymphocyte reduction is a clinically-validated marker of S1P_1_ desensitization, because in patients all S1P_1_ agonists tested to date were designed as S1P_1_-desensitizing agents to reduce peripheral blood lymphocytes and provide therapeutic benefit in MS.^33,34^ Consequently, the effect of selective S1P_1_ activation, without desensitization, on endothelial function in patients has not been studied to date. Rather, S1P_1_-desensitizing molecules, at doses characterized by marked S1P_1_ desensitization, demonstrate evidence of endothelial damage based on the incidence of macular edema, renal dysfunction and respiratory dysfunction in various patient populations.^33–38^

SAR247799 is a G-protein-biased S1P_1_ agonist (inhibiting adenylate cyclase preferentially over activation of β-arrestin and internalization pathways) with endothelial–protective properties in rats and pigs at doses that do not desensitize S1P_1_.^39^ Preclinical studies with SAR247799 showed protective effects in the renal and coronary vasculature in a preventative setting using single doses administered prior to an insult of ischemia/reperfusion-induced endothelial injury, but the effect of chronic administration of SAR247799 on the endothelium is currently unknown. Furthermore, to our knowledge, the effect of S1P_1_ activation on endothelial properties in patients has not been studied. In preclinical models, SAR247799 is bi-phasic molecule that can activate S1P_1_ with endothelial-protective properties at low doses, while causing lymphocyte reduction, like other S1P_1_- desensitizing molecules, at higher doses. Biological activities often follow the 4-parameter logistic model in which E_max_ is related to dose (i.e., the bigger the dose, the larger the effect),^40^ and with this assumption, many proof-of-concept trials test a single dose, at-or-close-to the maximum tolerated dose. However, given that S1P_1_ desensitization has been associated with endothelial-damaging effects, a U-shaped dose response of SAR247799 for endothelial-protective effects was considered pharmacologically plausible. Therefore, to reduce the likelihood of missing the clinical efficacy of the compound in patients, there is a need to understand the dose-response relationship of SAR247799 for endothelial-protective effects, under repeated-dosing conditions, and put this in the context of the level of S1P_1_ desensitization.

The aim of the current study was to determine whether 4-week therapy with SAR247799 improves reactivity of the dysfunction endothelium in type-2 diabetes patients using flow-mediated dilation (FMD). FMD is a technique that is regarded as a gold standard,^41^ predicts cardiovascular events,^42^ and can reveal pharmacological improvements in diabetes patients.^43–51^ We also compared the level of response to sildenafil; a positive control with established efficacy on FMD in this population.^43,45,46^ As SAR247799 is a biphasic molecule, we payed close attention to choosing doses that would be S1P_1_-activating, while limiting S1P_1_- desensitization, and performed a 5-week repeated-dosing study in a rat model of type-2 diabetes to facilitate the selection of appropriate doses for human testing. Zucker diabetic fatty (ZDF) rats^52^ have a defect in the leptin receptor gene (LEPR) and were used because they closely match the pathological characteristics of type-2 diabetes mellitus (T2DM) including hyperglycemia, insulin resistance, obesity and dyslipidemia.^53^ We first used the rat model to address whether chronic administration of SAR247799 has endothelial-protective properties and the level of S1P_1_ desensitization associated with such effects. The concentration-effect relationships of SAR247799 in the rat model were used to guide selection of doses for evaluation on endothelial function in type-2 diabetes patients.

## RESULTS

### Pharmacokinetics, lymphocyte and heart rate effects of SAR247799 in rats

To assess the ability of different dose strengths of SAR247799 to desensitize S1P_1_, peripheral blood lymphocyte counts were assessed following 5 weeks of oral administration to ZDF rats, alongside an assessment of the plasma concentrations of SAR247799.

The low and intermediate dose strengths of SAR247799 (0.002 and 0.007% (w/w) chow) did not reduce lymphocytes (Fig. 1A). The high dose strength (0.0245 % (w/w) chow) caused a 35% decrease in peripheral blood lymphocytes (Fig. 1A), indicating a partial desensitization of S1P_1_ at this dose. None of the doses of SAR247799 altered heart rate (Fig. 1B).

**Figure 1.**
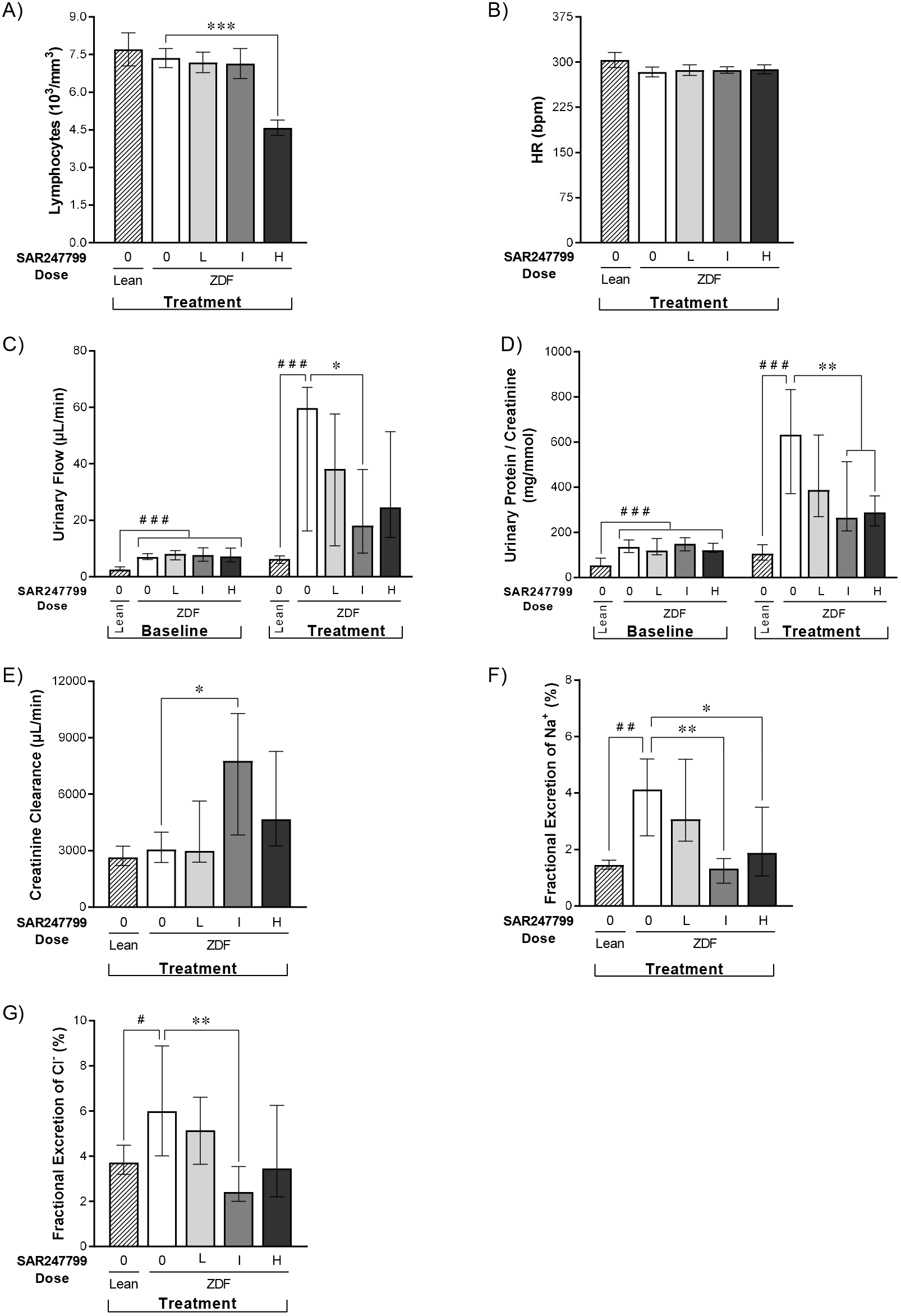
Effects of SAR247799 on lymphocytes, heart rate and renal parameters in diabetic rats. ZDF rats treated for 5 weeks with chow formulated with control (0), low (L; 0.002% w/w), intermediate (I; 0.007% w/w), or high (H; 0.0245% w/w) doses of SAR247799. Age-matched lean rats treated with control chow (0) for comparison. N=8, 12, 14, 12 rats in Lean, ZDF 0, ZDF L, ZDF I, ZDF H groups, respectively. Bars are mean ± SEM when Shapiro-Wilks test did not reject normality hypothesis (A, B) and median ± IQR otherwise (C, D, E, F, G). ^#^ p<0.05, ^##^ p<0.01, ^###^ p <0.001 for Lean vs ZDF control group; A and B by Student’s t test; E, F, G by Wilcoxon test; C and D baseline comparisons of Lean vs the four ZDF groups by Kruskal-Wallis followed by Wilcoxon post-hoc test; C and D treatment comparisons of Lean vs ZDF control group by Wilcoxon test. * p<0.05, ** p<0.01, *** p<0.001 for treated groups vs ZDF control group; A and B by one-way Anova followed by Dunnett’s test; E, F, G and C & D (treatment) by Kruskal-Wallis test followed by Wilcoxon post-hoc test.

Plasma concentrations of SAR247799 increased consistent with dose proportionality from low to intermediate dose strength, with mean AUC values of 2.27 and 10.2 μg.h/ml, respectively (Table 1). At the high dose strength there was a 10.5-fold increase in plasma concentration (106 μg.h/ml) for a 3.5-fold increase in dose from the intermediate dose.

**Table 1.**
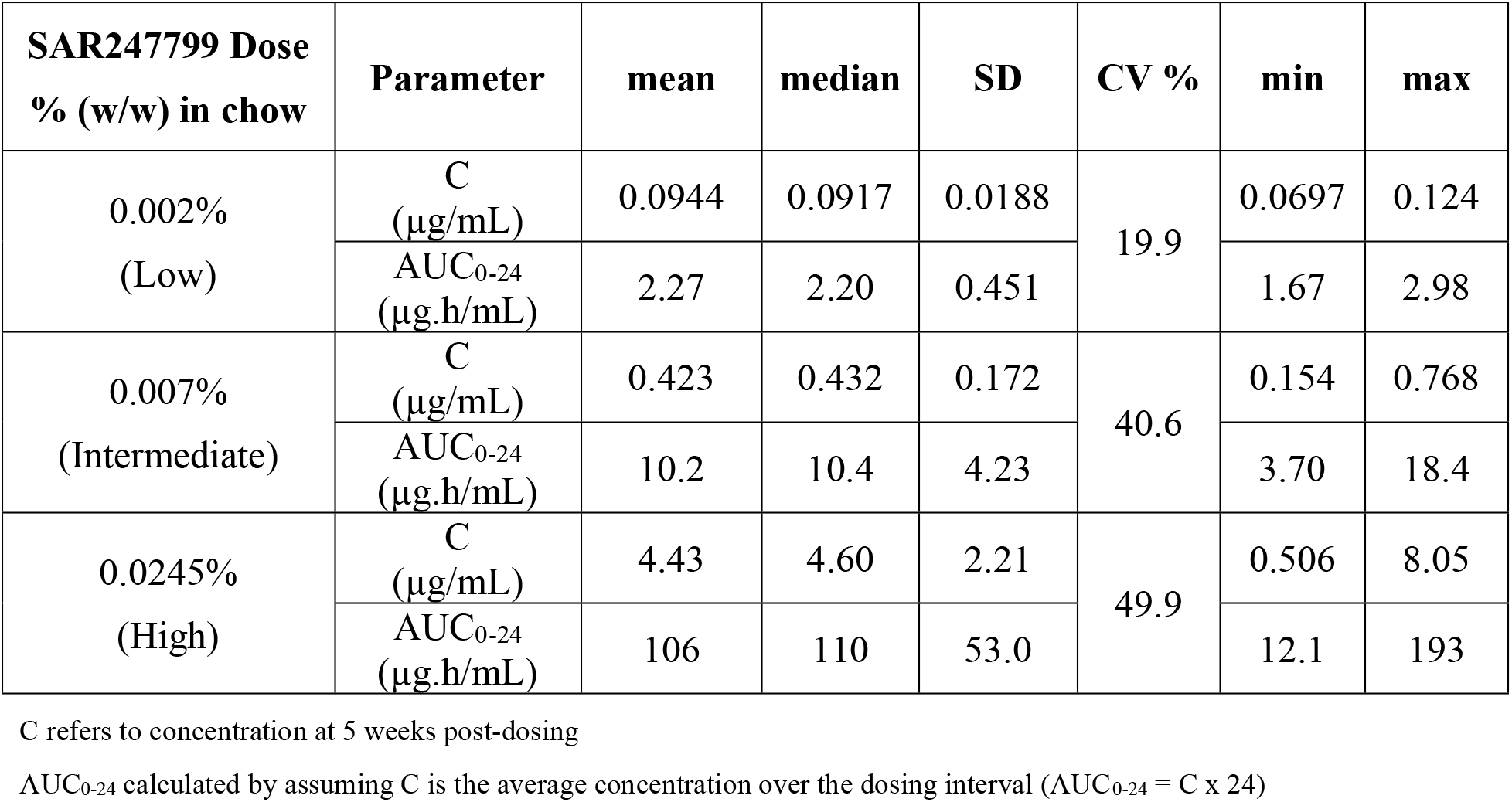
Descriptive statistics of plasma pharmacokinetics parameters in diabetic rats.

| **SAR247799 Dose**<br>**% (w/w) in chow** | **Parameter** | **mean** | **median** | **SD** | **CV %** | **min** | **max** |
| --- | --- | --- | --- | --- | --- | --- | --- |
| 0.002%<br>(Low) | C<br>($\mu\text{g/mL}$) | 0.0944 | 0.0917 | 0.0188 | 19.9 | 0.0697 | 0.124 |
| | AUC<sub>0-24</sub><br>($\mu\text{g.h/mL}$) | 2.27 | 2.20 | 0.451 | | 1.67 | 2.98 |
| 0.007%<br>(Intermediate) | C<br>($\mu\text{g/mL}$) | 0.423 | 0.432 | 0.172 | 40.6 | 0.154 | 0.768 |
| | AUC<sub>0-24</sub><br>($\mu\text{g.h/mL}$) | 10.2 | 10.4 | 4.23 | | 3.70 | 18.4 |
| 0.0245%<br>(High) | C<br>($\mu\text{g/mL}$) | 4.43 | 4.60 | 2.21 | 49.9 | 0.506 | 8.05 |
| | AUC<sub>0-24</sub><br>($\mu\text{g.h/mL}$) | 106 | 110 | 53.0 | | 12.1 | 193 |

C refers to concentration at 5 weeks post-dosing
AUC<sub>0-24</sub> calculated by assuming C is the average concentration over the dosing interval ( $\text{AUC}_{0-24} = C \times 24$ )

### SAR247799 improves renal function in diabetic rats independent of lymphocyte reduction

Compared to lean control rats, ZDF rats had higher body weight and higher plasma fructosamine levels (consistent with their diabetic status) (Supplementary Fig. IA, IB). 5-week treatment with SAR247799 (at any dose) did not alter fructosamine levels (Supplementary Fig. IB), consistent with SAR247799 not being a classical anti-diabetic drug controlling glucose levels. To determine the efficacy of SAR247799 following repeated administration, a variety of renal parameters were measured following 5-week SAR247799 treatment.

Urinary output was 10-fold higher in vehicle-treated ZDF rats compared to lean rats at the end of the 5-week treatment period (Fig. 1C). SAR247799 treatment significantly blunted the increase in urinary output at the intermediate dose (Fig. 1C).

The urinary protein: creatinine ratio (uPCR) was 6-fold higher in diabetic rats compared to lean controls (Fig. 1D). SAR247799 caused a dose-dependent reduction in uPCR, which was statistically significant at the intermediate and high doses. The lowest dose showed a partial effect and the top two doses caused similar effects, suggesting that a maximal PD effect on this parameter had been reached at the intermediate dose (Fig. 1D).

Creatinine clearance was similar in ZDF and lean rats (Fig. 1E). 5-week treatment with the intermediate dose of SAR247799 caused a 2.5-fold increase in creatinine clearance compared to vehicle-treated ZDF rats. There was no increase at the low dose, and a lesser (non-significant) increase at the high dose.

The fractional excretion of electrolytes (Na^+^ and Cl^−^) was increased in ZDF rats compared to lean (Fig. 1F, 1G). These increases were most effectively reduced by the intermediate dose of SAR247799, with levels similar to lean control rats, and to a lesser extent at the high dose.

Overall, the dose-response characterization on renal parameters (Fig. 1C-G) demonstrated that SAR247799 had maximal pharmacological effects on all parameters at the intermediate dose; a dose that did not reduce lymphocytes (Fig. 1A) or heart rate (Fig. 1B). There was a partial or lesser effect on most parameters at the low and high doses (Fig. 1C-G).

### SAR247799 improves endothelial biomarkers in diabetic rats independent of lymphocyte reduction

To assess the ability of repeated administration of SAR247799 to alter biomarkers of endothelial function the level of the following factors was assessed following 5-week treatment: vWF (coagulation), sICAM-1, sVCAM-1 and sE-selectin (adhesion molecule shedding), sPECAM-1 (capillary integrity), nitrite/nitrate and CRP (inflammation). All markers showed an increase in diabetic compared to lean rats and these achieved statistical significance except for vWF and nitrite/nitrate (Fig. 2A-G). SAR247799 treatment caused a dose-dependent reduction in all of these markers; the reductions were statistically significant for at least one dose for all biomarkers except sPECAM-1. Overall, SAR247799 showed the most effective reduction of endothelial biomarkers at the intermediate and high doses, with little difference between them.

**Figure 2.**
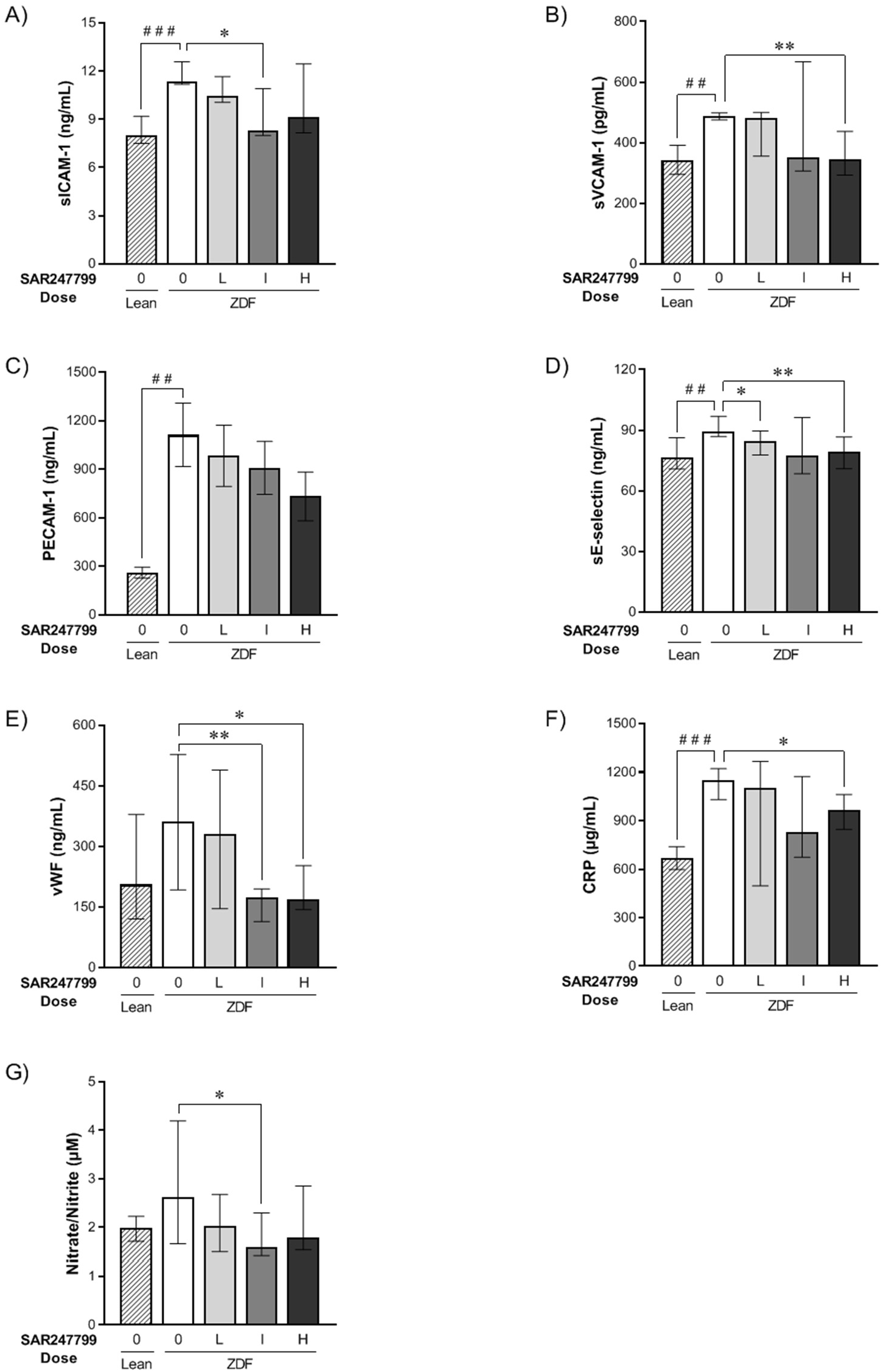
Effects of SAR247799 on plasma biomarkers in diabetic rats. ZDF rats following 5-week treatment with chow formulated with control (0), low (L; 0.002% w/w), intermediate (I; 0.007% w/w), or high (H; 0.0245% w/w) doses of SAR247799. Age-matched lean rats treated with control chow (0) for comparison. N = 8, 12, 14, 12 rats in Lean, ZDF 0, ZDF L, ZDF I, ZDF H groups, respectively. Bars represent mean ± SEM when Shapiro-Wilks test did not reject normality hypothesis (C) and median ± IQR otherwise (A, B, D, E, F, G).^##^ p<0.01,^###^ p<0.001 for Lean vs ZDF control group; Student’s t-test for C, or Wilcoxon test for A, B, D, E, F, G. * p<0.05, ** p<0.01, for treated groups vs ZDF control group; Kruskal-Wallis test followed by Wilcoxon post-hoc test for A, B, D, E, F, G.

Considering the combined renal function and plasma biomarker data (Fig. 1 and Fig. 2), the intermediate dose of SAR247799 provides the best overall efficacy profile. The lesser effects at the low dose could reflect sub-optimal exposure, and at the high dose could reflect partial S1P_1_ desensitization (35% reduction in lymphocytes) causing some loss of protective effect.

### Clinical Study population and safety assessments

We performed a randomized, double-blind, multi-center, active- and placebo-controlled clinical trial in type-2 diabetes patients to evaluate the activity of SAR247799 on endothelial function using flow-mediated dilation (FMD) (Fig. 3A). The baseline clinical characteristics are shown in Table 2. Patients progressed through the clinical trial as shown in Fig. 3B. Safety analysis included all 54 enrolled patients and is summarized in Table 3. There were no deaths, no serious or severe treatment-emergent AEs (TEAEs) and no AEs of special interest during the study. 53 of the total 54 patients enrolled in the study completed their assigned IMP for 28 days as planned. One patient, assigned to the sildenafil group, discontinued treatment on day 3 due to the occurrence of ventricular extrasystoles (grade 1 intensity according to NCI-CTCAE v4.03). Overall 19 of 54 patients experienced at least one TEAE during the study (Table 3), with the number of patients with TEAEs in the SAR247799 and sildenafil treatment groups being similar and slightly higher than in the placebo group. The most commonly reported TEAEs were headache and nasopharyngitis, with 5 and 4 patients reporting these TEAEs, respectively. The TEAEs were typically reported as mild, and all of them resolved with time.

**Figure 3.**
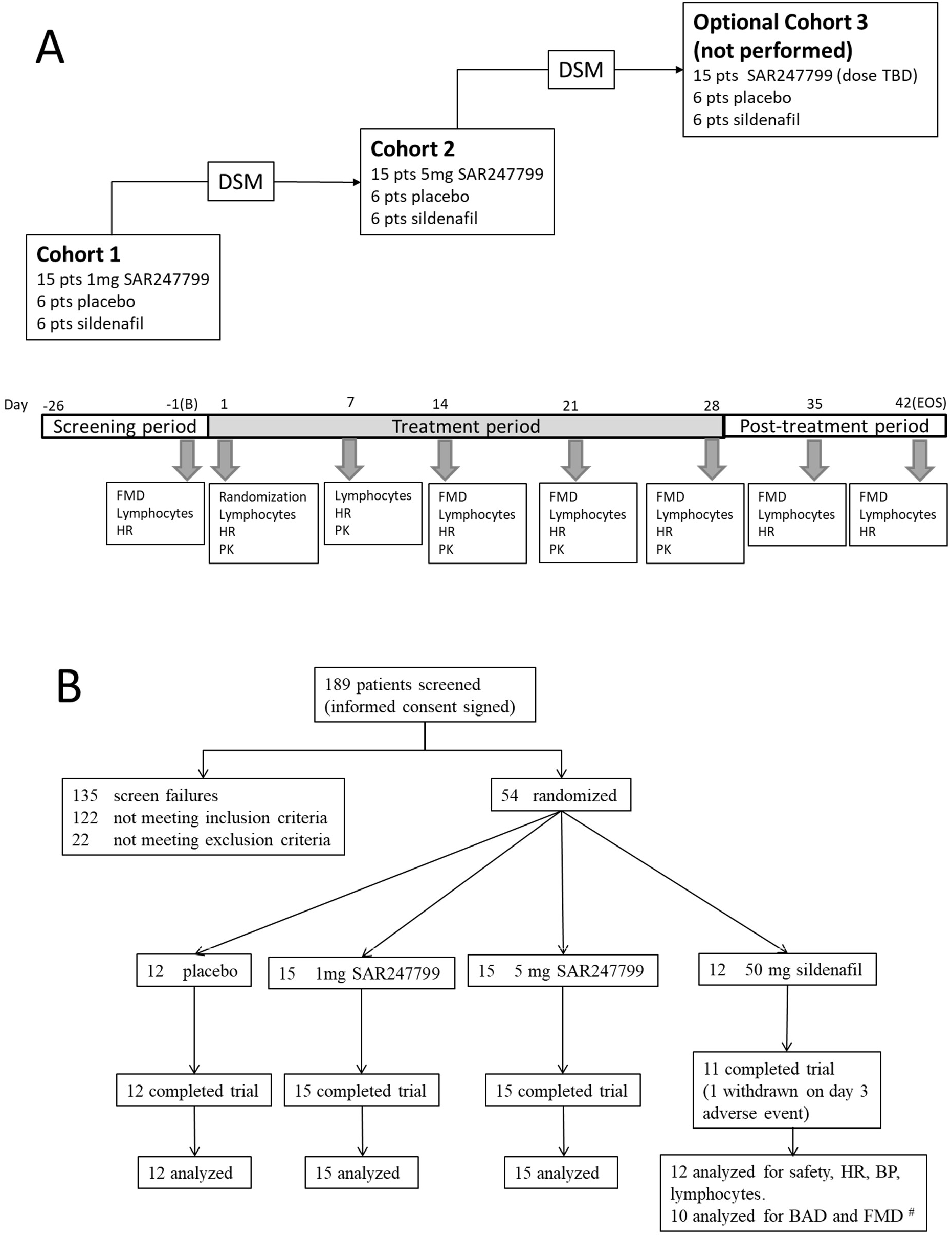
Design and patient progress through randomized, double-blinded clinical trial to evaluate SAR247799 on flow-mediated dilation in type-2 diabetes patients. A) Study design. B) Flow chart of patient progress through trial. # 1 patient excluded for unusable FMD data and 1 patient withdrawn. DSM, dose selection meeting; PK, pharmacokinetics; B, baseline; EOS, end of study

**Table 2.** Demographic and clinical variables at baseline.

| Variables |  | placebo<br>(N=12) | SAR247799 |  | sildenafil<br>(N=12) |
| --- | --- | --- | --- | --- | --- |
|  |  |  | 1 mg<br>(N=15) | 5 mg<br>(N=15) |  |
| Age in years | Mean (SD) | 56.3 (7.9) | 54.7 (7.0) | 56.4 (6.0) | 58.6 (3.8) |
| Gender | n females (%) | 3 (25) | 2 (13.3) | 6 (40) | 4 (33.3) |
| Race | [n (%)] |  |  |  |  |
| • White |  | 11 (91.7%) | 14 (93.3%) | 14 (93.3%) | 12 (100%) |
| • Black or African American |  | 0 | 1 (6.7%) | 1 (6.7%) | 0 |
| • Multiple |  | 1 (8.3%) | 0 | 0 | 0 |
| Ethnicity | n not hispanic or Latino (%) | 12 (100%) | 15 (100%) | 15 (100%) | 12 (100%) |
| Weight (kg) | Mean (SD) | 89.7 (13.1) | 87.9 (13.9) | 84.9 (13.1) | 88.3 (14.4) |
| BMI (kg/m<sup>2</sup>) | Mean (SD) | 29.9 (3.5) | 28.6 (3.0) | 29.1 (3.0) | 28.6 (3.7) |
| BMI | n <30 kg/m<sup>2</sup> (%) | 5 (41.7%) | 9 (60.0%) | 10 (66.7%) | 7 (58.3%) |
| HbA1c (%) | Mean (SD) | 7.02 (0.62) | 7.64 (0.68) | 6.84 (0.74) | 7.3 (0.63) |
| SBP (mmHg) | Mean (SD) | 133 (14) | 131 (12) | 123 (14) | (129 (12) |
| Heart rate (bpm) | Mean (SD) | 64 (7) | 69 (8) | 64 (6) | 68 (8) |
| FMD (%) | Mean (SD) | 3.88 (1.25) | 4.32 (1.67) | 3.87 (1.67) | 4.20 (1.52) |
| Concurrent therapy | N |  |  |  |  |
| • No treatment |  | 3 | 0 | 1 | 1 |
| • Oral anti-diabetic treatment |  | 8 | 13 | 14 | 11 |
| • ACEi/ARB |  | 4 | 4 | 3 | 7 |
| • Statins |  | 3 | 2 | 1 | 1 |
| • Thyroid replacement therapy |  | 1 | 1 | 4 | 0 |
| • Anti-steroidal anti-inflammatory |  | 1 | 1 | 1 | 1 |
| • Diuretic |  | 0 | 0 | 0 | 0 |
| • Other |  | 0 | 3 | 1 | 1 |

**Table 3.**
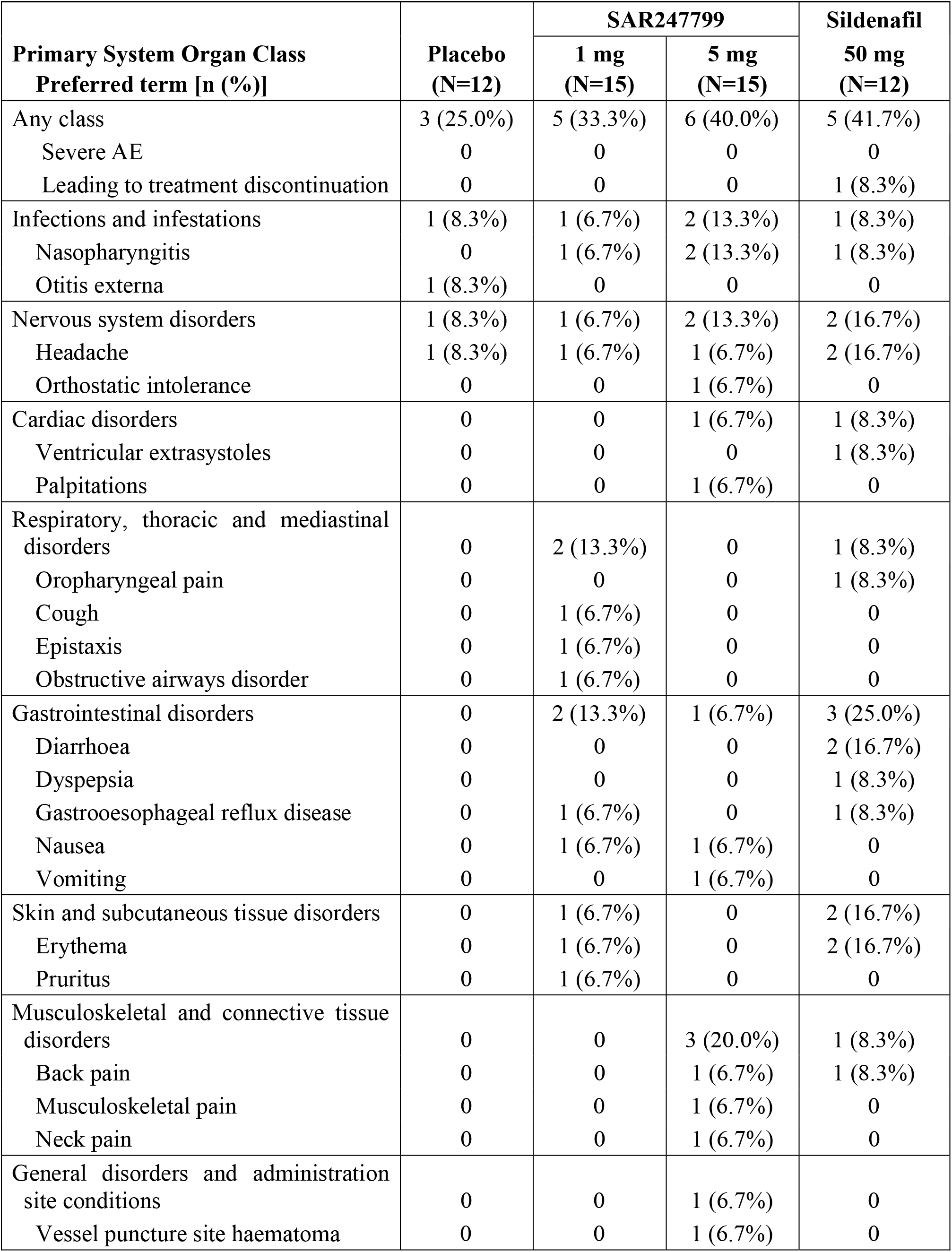
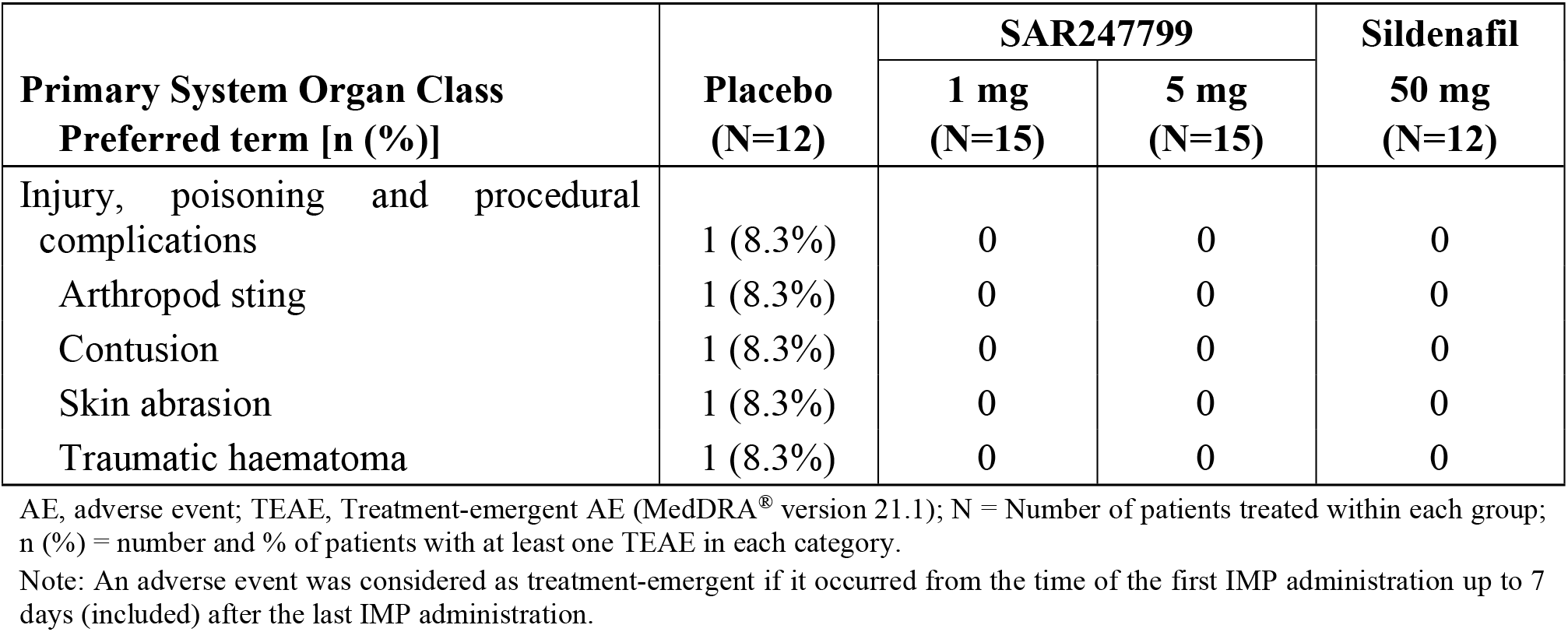
Number (%) of patients with TEAE(s) by primary system organ class and preferred term.

On day 1, SAR247799 caused a dose-dependent reduction in heart rate (mean reductions of 2 and 4 bpm at 1 and 5 mg, respectively) compared to no change in the placebo or sildenafil groups (Fig. 4A). As the study progressed (day 7–28), there was a mean 5–6 bpm heart rate increase in the placebo group, a similar (3–6 bpm) increase in the sildenafil group, a smaller (0–4 bpm) increase in the 5 mg group and no increase in the 1 mg SAR247799 group. Relative to the placebo group, the maximum heart rate decrease was observed with 5 mg SAR247799 on day-14 (mean difference versus placebo –6.4 bpm; 95% CI –2.1 to –10.7 bpm; p=0.004). Relative to the placebo group, in the 1 mg SAR247799 group a lower heart rate was sustained over the 28-day treatment period, but in the 5 mg SAR247799 group the heart rate effects diminished slightly on day 21 and day 28 (suggesting the onset of S1P_1_ desensitization).

**Figure 4.**
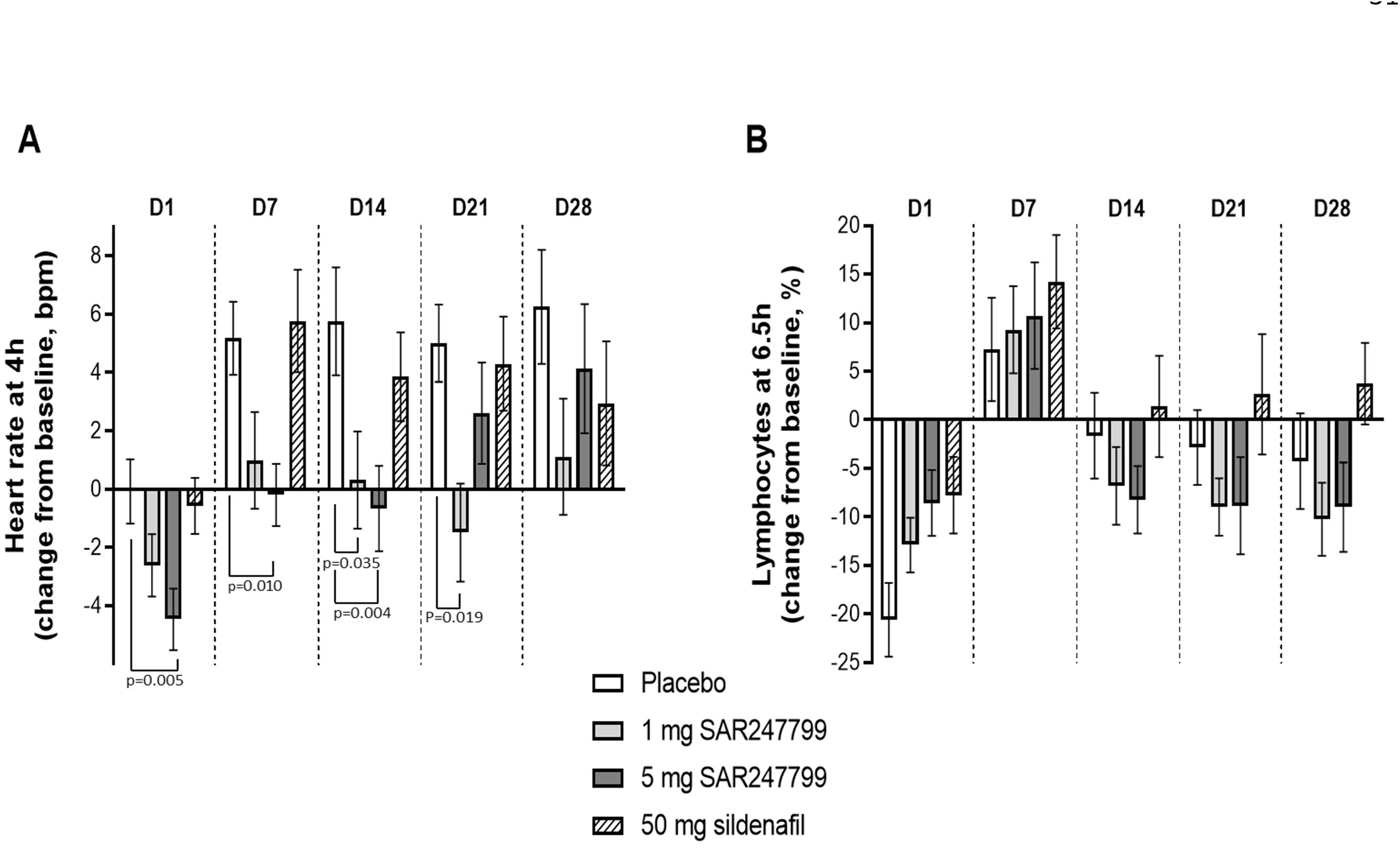
Effect of SAR247799 (1 and 5 mg), sildenafil (50 mg), or placebo on heart rate (A) and lymphocytes (B) in type-2 diabetes patients. Heart rate and lymphocytes expressed on each day (D1-D28) at time points associated with maximal pharmacodynamic effects (4 and 6.5 hours, respectively). Mean ± SEM; p values shown when <0.05 (repeated-measure model as described under Methods).

Systolic and diastolic blood pressure were slightly reduced over the treatment period, but without significant differences between placebo and treatment groups (Supplementary Fig. IIA, IIB).

### Pharmacokinetics and lymphocyte-reducing effects of SAR247799 in patients

To assess the ability of the two doses of SAR247799 to desensitize S1P_1_, peripheral blood lymphocyte counts were assessed over the 28 days of oral treatment to type-2 diabetes patients. The two doses of SAR247799 showed lymphocyte changes which were indistinguishable from placebo (Fig. 4B) (mean reductions versus placebo <8% at all timepoints).

Plasma concentrations of SAR247799 increased from day 1 to day 14 with an accumulation ratio of 2.5–2.9 and were consistent with dose proportionality between 1 and 5 mg/day (Table 4). The steady state AUC_0–24_h of SAR247799 were similar between the 1 mg/day human dose compared to the low dose in rat (2.97 μg/ml versus 2.27 μg/mL, respectively; Table 1 and Table 4). Similarly, the steady state AUC_0–24_h were similar between the 5 mg/day human dose and the intermediate dose in rat (14.9 μg/mL versus 10.2 μg/mL, respectively; Table 1 and Table 4). Both human doses (and the low and intermediate rat doses) did not significantly reduce lymphocytes (Fig. 1A and Fig. 4B). The AUC_0–24_h at the high dose in rats, inducing partial S1P_1_ desensitization (Fig. 1A), was 7.5-fold higher than the 5 mg/day human dose (106 μg.h/mL versus 14.9 μg.h/mL, respectively; Table 1 and Table 4).

**Table 4.** Descriptive statistics of plasma pharmacokinetic parameters in type-2 diabetes patients following 1.0 and 5.0 mg SAR247799 repeated once daily oral administration by dose level on Day 1 and Day 14.

| Day | Parameter | Dose | N | Mean | Median | SD | CV% | Min | Max |
| --- | --- | --- | --- | --- | --- | --- | --- | --- | --- |
| **1** | **T<sub>max</sub>**<br>(h) | 1 | 15 | 2.68 | 2.50 | 0.320 | 11.9 | 2.25 | 3.25 |
|  |  | 5 | 15 | 2.42 | 2.50 | 0.122 | 5.05 | 2.25 | 2.5 |
|  | **C<sub>max</sub>**<br>(µg/mL) | 1 | 15 | 0.0574 | 0.0538 | 0.0131 | 22.8 | 0.0403 | 0.0907 |
|  |  | 5 | 15 | 0.310 | 0.315 | 0.0635 | 20.5 | 0.207 | 0.414 |
|  | **AUC<sub>0-24</sub>**<br>(µg.h/mL) | 1 | 15 | 1.03 | 0.983 | 0.232 | 22.5 | 0.750 | 1.69 |
|  |  | 5 | 15 | 5.31 | 5.31 | 0.955 | 18.0 | 3.75 | 6.87 |
| **14** | **T<sub>max</sub>**<br>(h) | 1 | 15 | 2.24 | 2.25 | 0.0969 | 4.32 | 2.00 | 2.50 |
|  |  | 5 | 15 | 2.15 | 2.25 | 0.116 | 5.40 | 2.00 | 2.25 |
|  | **C<sub>max</sub>**<br>(µg/mL) | 1 | 15 | 0.152 | 0.147 | 0.0419 | 27.6 | 0.0998 | 0.271 |
|  |  | 5 | 15 | 0.779 | 0.760 | 0.186 | 23.9 | 0.500 | 1.13 |
|  | **AUC<sub>0-24</sub>**<br>(µg.h/mL) | 1 | 15 | 2.97 | 2.78 | 0.902 | 30.4 | 1.84 | 5.46 |
|  |  | 5 | 15 | 14.9 | 14.8 | 4.03 | 27.0 | 8.74 | 22.3 |
|  | **Rac_AUC<sub>0-24</sub>** | 1 | 15 | 2.86 | 2.70 | 0.432 | 15.1 | 2.10 | 3.56 |
|  |  | 5 | 15 | 2.79 | 2.73 | 0.492 | 17.6 | 2.10 | 3.64 |
|  | **Rac_C<sub>max</sub>** | 1 | 15 | 2.66 | 2.50 | 0.443 | 16.7 | 1.89 | 3.41 |
|  |  | 5 | 15 | 2.54 | 2.46 | 0.468 | 18.4 | 1.86 | 3.49 |

Rac, accumulation ratio.

### SAR247799 improves flow-mediated dilation in type-2 diabetes patients

52 of the 54 patients enrolled in the trial had FMD data that was analyzed (Fig. 3B). One patient in the sildenafil group was discontinued from treatment on day 3, as previously mentioned, and one further patient (also in the sildenafil group) was excluded from the FMD analysis because the FMD curves during hyperemia did not show the typical shape of the brachial diameter curve. At baseline, the mean FMD was well below the 7% threshold for inclusion into the study and ranged from 3.8% to 4.3% in the 4 treatment groups (Table 2). The pre-hyperemia BAD remained unchanged in all groups over the 42-day study, indicating that the arterial vasomotor tone was not affected and did not interfere with the FMD measurements (Supplemental Fig. IIC).

Variations in FMD were first assessed by comparing the mean placebo values over the 42-day study period. The placebo group showed mean increases compared to baseline of 0.7%, 0.6% and 0.7% on D14, D21 and D28 respectively, and minimal change at D35 and D42 (end of study) (Fig. 5A). Given this variation in the placebo group, FMD changes at each time-point were expressed as placebo-corrected values (Fig 5B).

**Figure 5.**
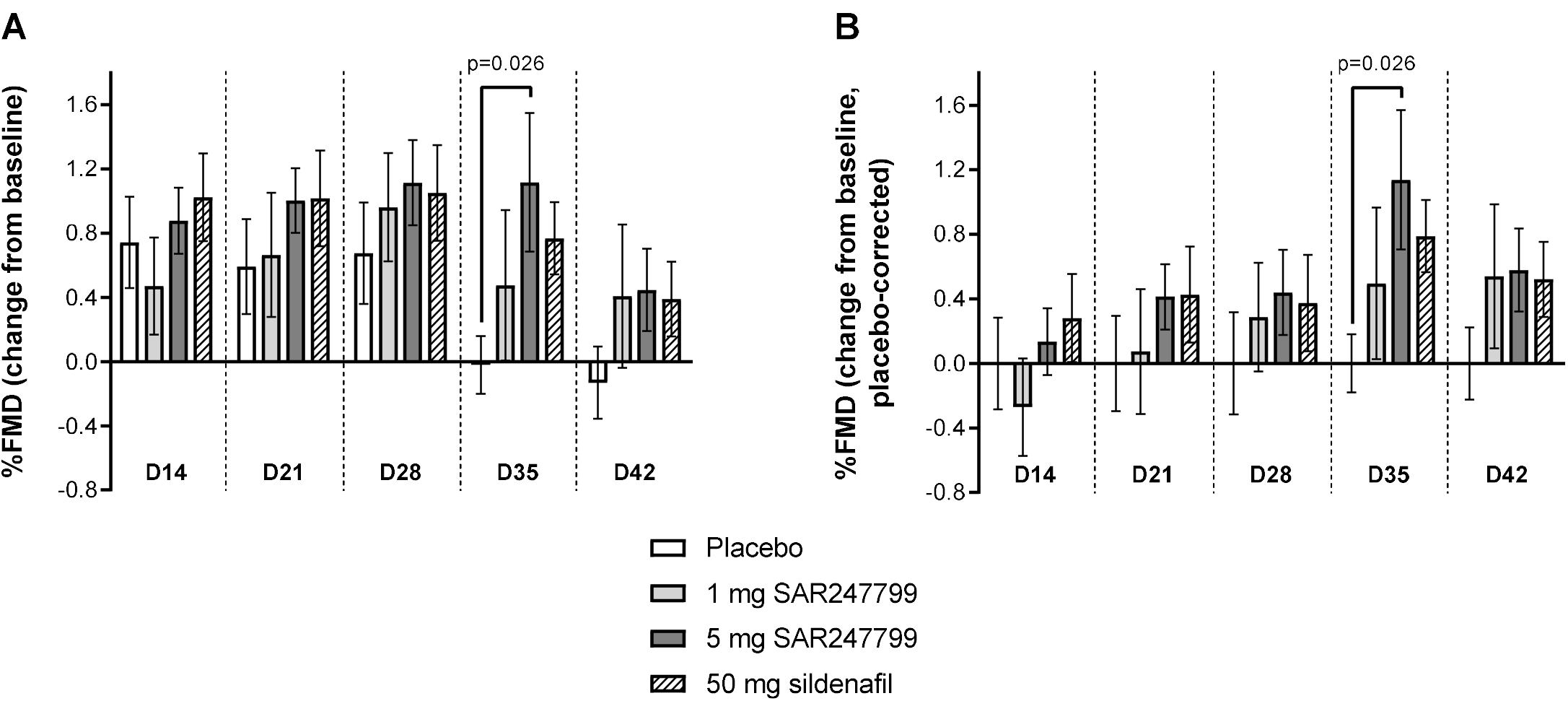
Effect of SAR247799 (1 and 5 mg), sildenafil (50 mg), or placebo on % flow-mediated dilation (FMD) in type-2 diabetes patients. FMD data expressed as change from baseline (A), or change from baseline, placebo corrected (B). Bars are mean ± SEM. Variation in the placebo groups in B (placebo-corrected FMD change) illustrated by retaining error bars with mean zero (first group at each time point). Precise p-values for difference versus placebo shown when p<0.05; repeated-measure model as described under Methods. Number of patients with FMD data in each histogram was as follows for placebo/ 1 mg SAR247799/ 5 mg SAR247799/ 50 mg sildenafil, respectively; 12/15/15/10 (D14, D21, D28 and D42) and 12/15/13/10 (D35).

The mean FMD change from baseline with 50 mg sildenafil (positive control) and 5 mg SAR247799 was higher than placebo at all time points up to D42 (Fig. 5A, Fig. 5B). For all three treatment groups (1 and 5 mg SAR247799 and 50 mg sildenafil) the maximum FMD change from baseline versus placebo occurred at 1 week after the end of treatment (D35), indicating that an effect had persisted after compound wash-out. On D35, mean differences versus placebo were 0.60% (95% CI –0.34% to 1.53%; p=0.203) for 1 mg SAR247799, 1.07% (95% CI 0.13% to 2.01%; p=0.026) for 5 mg SAR247799 and 0.88% (95% CI –0.15% to 1.91%; p=0.093) for 50 mg sildenafil. Overall SAR247799 showed a dose and time-dependent improvement in FMD, with effects at least as good as sildenafil.

## DISCUSSION

The study evaluated, for the first time, the pharmacological effect of S1P_1_ activation on endothelial function in patients. The experiments expand on a wealth of preclinical studies implicating S1P_1_ in regulating endothelial function and provide the first relevance of the S1P_1_ mechanism-of-action to patients with endothelial dysfunction. SAR247799 treatment progressively improved endothelial function (FMD) over 4 weeks in type-2 diabetes patients to levels that were in a similar range to sildenafil, and the vascular effects of SAR247799 were independent of lymphocyte reduction (unlike S1P_1_ functional antagonists used for multiple sclerosis treatment) or blood pressure-lowering.

SAR247799 showed a dose-dependent effect (5 mg > 1 mg > placebo) at each of the time-points evaluated, and the 5 mg dose was at least as effective as 50 mg sildenafil. An important finding of our study was that the effect of SAR247799 persisted after the end of treatment and compound wash-out, suggesting a potential disease-modifying effect to the endothelium. As the effect appeared to be both time- and dose-dependent, it is difficult to conclude whether the maximal possible effect of SAR247799 on FMD was reached. However, as SAR247799 was at least as effective as sildenafil under the conditions evaluated, a level of clinical relevance of the S1P_1_ mechanism has been established that warrants further evaluation in patients, including settings with longer treatment duration and/or with more relevant clinical endpoints.

Our results are consistent with a vascular function study performed in renal transplant recipients with 2.5 mg fingolimod.^38^ The 2.5 mg dose was 5-fold higher than the approved dose in multiple sclerosis patients (0.5 mg), caused marked S1P_1_ desensitization (70–80% lymphocyte reduction), and was associated with creatinine clearance impairment in renal transplant recipients.^37,38^ The study, performed 1.5 years post-transplant, demonstrated that FMD improved following 3-month fingolimod discontinuation. Although the investigators did not measure FMD prior to initiating fingolimod treatment, we hypothesize that the 2.5 mg dose (which was associated with marked S1P_1_-desensitization) caused endothelial damage, and the FMD improvement seen over the 3-month fingolimod-withdrawal period reflects a restoration of endothelial function. In combination with the findings from the current study, S1P_1_ activation and desensitization appear to improve and impair FMD, respectively.

PDE5 inhibitors decrease forearm vascular resistance by direct vascular smooth muscle effects (e.g. stimulating guanylate cyclase) as well as endothelium-dependent effects (release of vasodilatory factors including nitric oxide). High dose (100 mg) sildenafil failed to improve FMD in type-2 diabetes patients, but caused marked vasodilation as evidenced by blood pressure-lowering and dilation of the BAD.^55^ Two studies with lower doses of sildenafil (50 mg q.d. and 25 mg t.i.d.) showed 5–6% increases from baseline FMD values in the 7–9% range, and with no evidence of vasodilation.^43,45^ Thus, we selected 50 mg q.d. sildenafil for the current study, confirmed no vasodilatory effect (i.e., no blood pressure-lowering or BAD increase) and showed an increase in FMD from 4% at baseline to about 5% with treatment. While it is not immediately apparent why sildenafil showed a smaller effect on FMD in our study, it is possible that the enrichment of patients for marked endothelial dysfunction in our study (lower baseline FMD) could have influenced the absolute effect of sildenafil. Nevertheless, the inclusion of sildenafil as positive control allowed us to appropriately benchmark the effect of SAR247799 to a clinical calibrator and show that this new S1P_1_ mechanism-of-action can produce effects on FMD at least as effective as PDE5 inhibition.

The improvement in FMD demonstrated with SAR247799 may have clinical implications for cardiovascular (CV) risk reduction. FMD, in addition to an index of endothelial dysfunction, predicts the rate of progression of carotid intima media thickness (CIMT) with superiority to the Framingham score.^56^ Furthermore, drugs that improve FMD have proved beneficial on CV outcomes such as statins and anti-hypertensive drugs (including ACE-Inhibitors, angiotensin I receptor blockers, and calcium channel blockers) with average effects for these drug classes of 1–1.5% increase in FMD.^49–51,57,58^ Over a broad range of populations, including patients with CV disease, a 1% improvement of FMD is associated with a 12% reduction in CV events.^42^ As noted above, since FMD improvement with sildenafil varied between studies, it is difficult to extrapolate quantitatively the benefit of SAR247799 on CV risk reduction based on the existing data alone. SAR247799 is also expected to improve the microvasculature (as shown in animal models) and indices of microvascular function predict CV risk independent of FMD.^42,59^ The potential CV risk reduction with SAR247799 could be underestimated by relying on the FMD (macrovascular) contribution alone, and further studies to assess microvascular function are needed.

All previous S1P_1_ functional antagonists tested in humans cause first dose heart rate reduction, which is time-dependently lost as S1P_1_ desensitization sets in. In our clinical study, 1 mg SAR247799 caused a small and sustained level of heart rate reduction over 28 days compared to placebo, whereas the 5 mg dose showed some evidence for desensitization on D21 and D28. While heart rate reduction and endothelial protection are both activities associated with S1P_1_ activation, they exhibited different kinetics: heart rate reduction displayed a rapid onset-of-action and was subject to rapid desensitization, whereas endothelial vasoreactivity (FMD) displayed a progressive time-dependent effect with propensity for desensitization that is not yet characterized. As SAR247799 distributes with higher concentrations in plasma compared to heart,^39^ the demonstration of sustained cardiac pharmacology over 28 days with 1 mg SAR247799 suggests that doses ≤1 mg may warrant evaluation for endothelial effects in studies of longer duration.

Although the long-term CV safety profile of SAR247799 remains to be established, it has been suggested that FMD trials play an important role in de-risking new CV agents. For example, the first CETP inhibitor, torcetrapib, unexpectedly demonstrated an increase in CV events in the > 15,000 patient ILLUMINATE trial, despite a substantial increase in HDL cholesterol, and this adverse signal was subsequently associated with a pharmacological decrease in FMD.^60,61^ In light of a subsequent CETP inhibitor, dalcetrapib, being neutral on both FMD and CV events,^22,62,63^ it has been proposed that demonstrating a lack of adverse vascular effects or even vascular benefit through increasing FMD, as demonstrated here with SAR247799, increases confidence in the use of new treatments in large-scale clinical trials over years.^64^ The point is particularly noteworthy given that HDL is the main carrier of S1P in plasma, and there is correspondingly an elusive link between HDL-raising agents and S1P_1_ agonists.

Our experiments were translational in nature as we used two complementary experimental systems (human and rat) to study the endothelial properties of SAR247799 in the setting of diabetes. Since SAR247799 is a biphasic molecule, it was necessary to study doses providing sufficient S1P_1_ activation to cause endothelial-protective effects, while avoiding higher endothelial-damaging doses by not exceeding a certain threshold of S1P_1_ desensitization. The precise threshold of S1P_1_ desensitization (based on lymphocyte reduction) associated with unwanted endothelial properties is currently unknown. The preclinical dose-ranging provided the foundation for selecting two doses of SAR247799 that provided partial and maximal efficacy (based on renal function and endothelial biomarkers in rats), while avoiding a higher dose that was less efficacious and associated with a certain level of S1P_1_ desensitization (35% lymphocyte reduction). In diabetes patients, we observed that the two corresponding doses (1 and 5 mg, which yielded exposures that approximated the lower two animal doses), produced a similar dose-dependent endothelial effect (FMD change at 5 mg > 1 mg >placebo), while also avoiding lymphocyte reduction. As there was a time-dependent FMD increase with SAR247799 over 35 days, it would be important in subsequent studies to consider whether low doses (1 mg or less) might produce more profound effects with longer treatment duration. As higher doses or longer treatment times were not evaluated in patients, we cannot conclude whether the maximal endothelial effect for this mechanism of action had been reached.

The preclinical studies complement the clinical study with respect to the mechanism of action of SAR247799. The preclinical results suggest that SAR247799 could also improve renal function in patients (based on improvements in protein: creatinine ratio, creatinine clearance and fractional excretion of electrolytes) and are particularly relevant given the matching concentration-effect relationships between clinical (on FMD) and preclinical studies (on renal function and biomarkers). The creatinine clearance improvement demonstrated with S1P_1_-activating doses of SAR247799 in rats are consistent with a creatinine clearance decline with S1P_1_- desensitizing doses of fingolimod in renal transplant patients.^37^ Given the endothelial effects of SAR247799 demonstrated herein in humans, SAR247799 is an attractive drug candidate for evaluation in renal, cardiac and other manifestations of endothelial dysfunction.

The study had a number of limitations. Firstly, the FMD study could have benefited from a slightly larger sample size, especially as the number of patients in the placebo group ended up being imbalanced due to the optional third cohort not being conducted, and the p-value at day 35 with 5 mg SAR247799 (0.026) slightly exceeded our significance threshold that considered multiple-arm adjustment (α=0.025). Nevertheless, the inclusion of a positive control (sildenafil) that had demonstrated improvements in FMD in type-2 diabetes patients, facilitated interpreting the effects of SAR247799 in relation to a clinical benchmark. Secondly, given the finding of a time-dependent effect of SAR247799 on FMD, in hindsight, 4-week treatment duration may not have been optimal to characterize the maximal effects of this new mechanism of action on endothelial properties. Thirdly, we did not assess the direct vascular smooth muscle effects of the compounds by performing an endothelium-independent vasodilation test using sublingual nitroglycerin administration (due to its contraindication with sildenafil). However, in none of the treatment groups was blood pressure or BAD (pre-hyperemia) altered and, together with a lack of functional S1P_1_ receptors on vascular smooth muscle,^65^ our FMD results are unlikely to be influenced by a direct action on vascular smooth muscle.

Overall, these preclinical and clinical pharmacology findings in experimental models of type-2 diabetes demonstrate consistent endothelial properties of 4-to-5-week SAR247799 treatment. Endothelial effects were observed without lymphocyte reduction and at similar compound exposures between the human and animal systems. Given that SAR247799 improved FMD with similar effects to sildenafil under the conditions evaluated, and that SAR247799 did not lower blood pressure, the results support further evaluation of S1P_1_ agonists in diseases associated with endothelial dysfunction.

## MATERIALS AND METHODS

### Animal studies

All animal studies were performed in accordance with the European Community standard on the care and use of laboratory animals and approved by the IACUC of Sanofi R&D.

#### Animals

Five-week-old (≈125–150 g) male Zucker Diabetic Fatty Fa/Fa (ZDF) rats and Zucker Lean Fa/+ rats (Lean) (Charles River, France) were housed in groups of two in 1500 UTemp cages (Tecniplast, France) in 29/12 Plus sawdust (Souralit) enriched with aspen bricks and play tunnels (Datesand). They were kept in a climate-controlled environment (21–22°C, 55% humidity) with a 12:12 h light-dark cycle and provided with diet and water (0.2 µm filtered tap water) *ad libitum* throughout the duration of the study. Control diet A04 (SAFE) was used from the arrival of the animals up to 9 weeks of age.

#### Experimental Design

At 9 weeks of age, ZDF rats were randomized by body weight to a chow formulation: control chow (SSNIFF Spezialdiäten GmbH) or chow formulated with one of 3 dose strengths of SAR247799: low, intermediate and high corresponding to 0.002, 0.007 and 0.0245 % (w/w), respectively. 12 ZDF rats were included in each of the control, low and high dose groups. A slightly higher number of ZDF rats (14) were included in the intermediate dose group, as it was predicted to be the most effective dose. 8 Lean rats, fed with control chow, were included in the study to confirm the pathological status of ZDF rats (and the lower number was based on the principle of reduction of 3Rs). All analyses were performed by a blinded experimenter.

#### Urinary evaluation

18-hour urine samples were collected at baseline and after 5 weeks of treatment using metabolic cages (Tecniplast, France). Urinary protein, creatinine, chloride and sodium concentration were measured by ABX PENTRA 400 (Horiba). Creatinine clearance was calculated as urinary creatinine X urine volume/plasma creatinine and expressed as microliters per minute.

#### Plasma Assessments

Blood samples were collected in K2-EDTA tubes from the caudal vein under 1.5–2 % isoflurane anesthesia at baseline and after 5 weeks of treatment. Blood lymphocyte count was measured by MS9–5 (Melet Schloesing) in freshly collected blood, then blood samples were immediately centrifuged at 4°C/10 min/10,000g and the plasma was stored at –80 °C. Plasma creatinine and fructosamine were measured by ABX PENTRA 400 (Horiba). SAR247799 concentrations were measured from 5-week plasma samples using liquid chromatography tandem mass spectrometry.

Plasma biomarkers were measured by ELISA following 5 weeks of treatment according to manufacturers’ instructions. Soluble ICAM-1, sE-Selectin and von Willebrand Factor (vWF) were measured using MILLIPLEX MAP Rat Vascular Injury Magnetic Bead Panel 2 (Merck), sVCAM-1 from VCAM-1 Rat ProcartaPlex™ Simplex Kit (ThermoFisher Scientific), CRP from CRP Rat ProcartaPlex™ Simplex Kit (ThermoFisher Scientific) and sPECAM-1 from Rat PECAM-1 Elisa kit (MyoBioSource). Nitrate/nitrite concentration was measured by nitrate/nitrite fluorometric assay kit (Cayman Chemical) in plasma samples previously filtered with Microcon-10 kDa Centrifugal Filter Unit with Ultracel-10 membrane (Merck).

#### Heart Rate

Heart rate was measured at the end of the study under anesthesia (1.5–2% isoflurane) after all other assessments were completed. Heart rate was measured from consecutive R-R intervals in the ECG signals recorded from suitable electrodes connected to Vivid E9 (GE).

#### Statistical Analysis

SAS® version 9.2 for Windows 7 was used for statistical analysis. Single comparison was performed by 2- tailed unpaired Student’s t-test or Wilcoxon test, when normality was not met. Multiple comparisons were analyzed by one-way Anova, following by Dunnett’s post-hoc test in normally distributed and homogeneous parameters, otherwise a Kruskal-Wallis test followed by Wilcoxon two-tailed comparison test was performed. A p value of ≤0.05 was considered significant. Data are expressed as mean ± SEM for normally-distributed data, otherwise as median ± interquartile range (IQR).

### Clinical Study

The clinical study was conducted as a multi-center study by Profil (2 centers in Germany: Profil Institut für Stoffwechselforschung GmbH, Neuss and Profil Mainz GmbH & Co. KG, Mainz) and was approved by the competent authority as well as two independent ethics committees (Local Ethical Committee of Düsseldorf and Local Ethical Committee of Mainz, Germany). The study was performed in accordance with the clinical trial protocol and with ethical principles having their origin in the Declaration of Helsinki. All patients provided written informed consent before participating. The trial was prospectively registered with Unique identifier NCT0346201 on 12/03/2018 (https://www.clinicaltrials.gov). The study dosing scheme is summarized in Fig. 3A.

#### Patients

The study population included male and female type-2 diabetes patients, with > 6 months from diagnosis of diabetes at the time of screening, and otherwise healthy as assessed by clinical and laboratory assessments and detailed medical history. Main inclusion criteria included age 18–64, body mass index (BMI) 18–35 kg/m^2^, Hemoglobin A1c (HbA1c) <8.5%, flow-mediated dilation ≤7%, estimated glomerular filtration rate (eGFR) >60 mL/min/1.73 m^2^ and vital signs after 10 minutes rest in supine position in the following ranges: systolic blood pressure 95 to 160 mmHg, diastolic blood pressure 45 to 100 mmHg, heart rate 60 to 100 bpm. In addition, clinical laboratory evaluations and ECG recording were to be normal or without any clinically-relevant findings

Patients with reported pathologies (apart from stable T2DM status) which, as judged by the investigator, may affect the patient’s health, participation in, or the outcome of the study, were excluded. Particularly, any history of clinically-relevant cardiovascular disease (including established atherosclerosis or non-controlled hypertension) or pulmonary, renal, hepatic, ophtalmologic or infectious diseases led to exclusion. A few co-morbidities such as controlled dyslipidemia, controlled hypertension, stable and treated hypothyroidism were accepted.

Any medication which had a potential to interfere with safety, pharmacokinetics (PK), or pharmacodynamics (PD) of SAR247799 and sildenafil or study measurements was prohibited; this included nitrates, all calcium channel blockers (CCBs), phosphodiesterase type 5 (PDE5) inhibitors, guanylate cyclase (GC) stimulators, beta blockers, glucagon-like peptide-1 (GLP-1) agonists, insulins (all types), drugs which decrease heart rate, anticoagulants and antithrombotics (except aspirin), antiarhythmics (such as digoxin), recent (≤3 months) use of systemic immunosuppressive or corticosteroid therapy, any inactivated vaccination during study treatment, any attenuated vaccination within 2 months before inclusion, and any biologics (antibody or its derivatives) given within 4 months before inclusion. The following medications were authorized if patients were on stable therapy: oral antidiabetics other than GLP-1 agonists, angiotensin converting enzyme (ACE) inhibitors, angiotensin II receptor blockers (ARBs), statins, aspirin as antithrombotic therapy, diuretics, hormonal contraception, menopausal hormone replacement therapy, and thyroid hormone replacement therapy. Only patients smoking less than 5 cigarettes or equivalent per week were enrolled. Smoking and tobacco were not allowed during institutionalization periods.

A patient could be withdrawn from the study at any time for any of the following reasons: refusal to comply with the parameters of the study, desire to withdraw; medical condition which the investigator considered to contraindicate the continuation of the study; or the occurrence of an adverse event.

The above patient characteristics for the trial were modelled from previously performed FMD studies.^43–48^

#### Clinical Study design

The selected design was a double-blinded, double-dummy, randomized, placebo and active-controlled study. Such design was selected to allow comparison of SAR247799 to placebo as well as the positive control, sildenafil, which has been shown to improve FMD in a variety of populations including patients with type-2 diabetes.^43,45,46^ The study was performed with 27 patients per cohort (6 placebo, 6 sildenafil, and 15 on a given dose of SAR247799) who received the treatments for 28 days in a once-a-day regimen. Two cohorts with escalating doses of SAR247799 (1 and 5 mg) were conducted and the protocol had the option for a third cohort, which was not performed. Capsules, blisters and boxes of investigational medicinal product (IMP) (SAR247799, sildenafil (50 mg) or placebo) were indistinguishable to preserve the double-blind.

A decision to proceed from one dose to the next was made jointly by the sponsor and the investigator on the basis of reviewing the preliminary safety and PK exposure data of the previous dose.

The study comprised of a screening period of up to 26 days during which an FMD threshold of <7% was used to enrich for patients with dysfunctional endothelium. SAR247799, sildenafil and placebo capsules were taken under fasted conditions each morning from day 1 to day 28 (inclusive) and there was a 14-day follow-up period till the end of study visit on day 42. There was a 4-day in-house period (day –2 to day 3) followed by ambulatory conditions with various in-house periods for FMD assessments (day 13–14, day 20–21, day 27–28, day 34–35 and day 41–42).

Patients were randomized on day 1 to receive SAR247799, placebo or sildenafil in 5:2:2 ratio if they met all inclusion/exclusion criteria (at day –1 and/or day –2). Sanofi controlled the double-blind key for the study. The IMP was administered under medical supervision during in-house periods and patients were instructed to take the IMP under fasted conditions each morning during ambulatory periods for which compliance was checked by counting the number of returned capsules at the end of the study.

#### Endothelial function Assessments by flow-mediated dilation

Endothelial function was assessed by measuring FMD of the brachial artery following reactive hyperemia. Measurements were made during screening, at baseline (day –1) and on day 14, 21, 28, 35 and 42 (Fig. 1) and the same arm (right side when possible) was used throughout. Patients were institutionalized overnight prior to each assessment, during which smoking and tobacco were prohibited, and measurements made in the morning under fasted conditions. Patients were examined in a quiet temperature-controlled room and asked to avoid caffeinated beverages for 12 h prior to each assessment. The brachial artery diameter (BAD) was assessed by high-resolution ultrasound imaging above the elbow, initially after 15 min rest in a supine position, and then following reactive hyperemia which was induced by 5 minutes inflation of a pneumatic tourniquet inflated to a pressure of 200 mmHg around the forearm and sudden cuff-deflation. The focus zone was set to the depth of the anterior vessel wall and depth and gain settings were optimized to identify the lumen vessel wall interface. The maximum brachial artery diameter was determined by continuously imaging from prior to cuff inflation until 2 min after cuff release. The ultrasound images were recorded directly onto the hard disk of the ultrasound machine and image analysis performed using an automated edge-detection software (Vascular Research Tools 5, Medical Imaging Applications LLC, Coralville, Iowa, USA) as previously described.^54^ FMD was expressed as the percentage change in BAD following reactive hyperemia.

#### Other assessments

Lymphocytes and heart rate were assessed at timepoints as shown in Figure 1. Other safety assessments were performed throughout the study period and included adverse events (AE) recording, physical examinations, vital signs, laboratory tests (hematology, urinalysis, biochemistry including high sensitivity c-reactive protein [hsCRP]) and continuous ECG telemetry.

#### Pharmacokinetics

SAR247799 concentrations were measured from plasma samples at the following time points: 4 and 12 h post-dose on day 1, 0 h (pre-dose) on days 2, 3, 7, and at 3 h post-dose on days 14, 21, and 28. A validated liquid chromatography tandem mass spectrometry method with a lower limit of quantification (LLOQ) of 5 ng/mL was used. The following PK parameters were calculated with a Bayesian approach using a population PK model developed for SAR247799 in healthy subjects following single and repeated administration; maximum concentration observed (C_max_), time to reach C_max_ (T_max_), area under plasma concentration versus time curve calculated using a trapezoid method over 24 hours (AUC_0–24_) and accumulation ratio (Rac [Cmax, AUC_0–24_]).

#### Sample size considerations

Sample size calculations were based on previous FMD studies in type-2 diabetes patients where sildenafil produced at least a 3% FMD increase from baseline compared to placebo,^43,45,46^ and a standard deviation (SD) of 2.5% was assumed (corresponding to a statistical effect size of at least 1.2).

The sample size was calculated by assuming that 3 doses of SAR247799 would be evaluated over 3 cohorts. In order to control the global type I error rate, a one-sided α=1.67% (5% divided by 3 doses) was used. A sample size of 13 evaluable patients per group would have had 80% power to detect an increase of 3% of the change in FMD from baseline of SAR247799 versus placebo (assuming a common SD of 2.5%) with a one-sided 1.67% significance level. Considering that 10% of patients may not have evaluable baseline and post-baseline FMD, 15 included patients per group was considered necessary.

As the third cohort was optional, to permit the analysis at the end of two cohorts with one-sided α=2.5% and at least 80% power, the sample size for sildenafil and placebo was increased by one patient per cohort (from 5 to 6), resulting in the following distribution of patients randomized across the first 2 cohorts: 15 patients on 1 mg SAR247799, 15 patients on 5 mg SAR247799, 12 patients on placebo and 12 patients on sildenafil.

#### Statistical analysis

SAS® version 9.4 for Windows 7 was used for statistical analysis. Demographic data were summarized in descriptive statistics by treatment group. All biomarkers were also summarized in descriptive statistics by treatment group and time.

Count and percentage of patients experiencing an adverse event were provided for each treatment by Medical Dictionary for Regulatory Activities (MedDRA^®^ version 21.1) primary SOC and preferred terms.

FMD change from baseline on D14, D21, D28, D35 and D42 was analyzed within a repeated-measures model with fixed terms for treatment, time, treatment-by-time interaction and with gender and baseline value as covariate and with subject as a random term with an unstructured variance-covariance matrix. Mean difference between treatments and their corresponding 95% confidence interval were estimated within the model framework at each time.

Similarly, heart rate (change from baseline at 4 h) and lymphocytes (% change from baseline at 6.5 h) on D1, D7, D14, D21 and D28 were analyzed within the same repeated measures model as for FMD with mean difference between treatments and their corresponding 95% confidence interval estimated within the model framework at each time.

## Data Availability

All relevant preclinical and clinical data are provided within the paper and its supporting information files. Qualified researchers may request access to patient level data and related study documents including the clinical study report, blank case report form, statistical analysis plan, and dataset specifications. Patient level data will be anonymized, and study documents will be redacted to protect the privacy of trial participants. Further details on Sanofi’s data sharing criteria, eligible studies, and process for requesting access can be found at www.clinicalstudydatarequest.com.

## ACKNOWLEDGEMENTS

The authors thank the study participants, the clinical research staff at the 2 sites, Charlene Buffat and Thomas Rizard for trial management support, Ariane-Saskia Ries and Katharina Iwaniuk for study organization at Mainz and Neuss sites, respectively, Carol Damon for data programming, Anne Charlier, Clementine Chopin and Mélanie Mellerin for compound supply, Delphine Marin and Mélodie Hermier for data management, Michel Scemama for pharmacovigilance support, Helene Joyeux, Christine Maestrini and Tadashi Sugihara for regulatory support, Didier Simard and Brigitte Molinier for biomarker evaluation, Catherine Cadrouvele for *in vivo* study support, and Dorothee Tamarelle for certifying preclinical statistics.

## AUTHOR CONTRIBUTIONS

**Conceptualization**: LB, GA, LP, BP, RS, CH, DR, PJ, AJM, SK, PD, AAP. **Data curation**: MFE, BP, RB, XB. **Formal analysis**: LB, MFE, BP, OV, FH, RB, XB, DR, LH, AAP. **Funding acquisition**: BP, PJ, AJM, PD, AAP. **Investigation**: GA, LP, MFE, TH. **Methodology**: LB, GA, LP, MFE, BP, TH, JA, RS, CH, OV, FH, DR, AJM, LH, SK, PD, AAP. **Project administration**: AT, JA. **Resources**: GA, LP, AT, JA. **Supervision**: LB, GA, LP, BP, PJ, AJM, LH, SK, PD, AAP. **Validation**: GA, LP, BP, LB, AAP. **Visualization**: MFE, BP, XB, AAP. **Writing – original draft**: LB, MFE, BP, AAP. **Writing – review and editing**: GA, LP, AT, ME, TH, JA, HC, RS, CH, OV, FH, RB, XB, DR, PJ, AJM, LH, SK, PD. **Accountable for accuracy, integrity and approval of final version**: all authors.

## ADDITIONAL INFORMATION

### Funding

This work was supported by Sanofi.

### Competing interests

At the time of conduct of the studies, LB, MFE, BP, AT, OV, FH, RB, XB, DR, PJ, AJM, LH, SK, PD and AAP were employees of Sanofi, GA, LP, ME, TH, JA and HC were employees of Profil Institute, RS and CH received personal fees from Profil Institute. Authors affiliated with Sanofi may have equity interest in Sanofi. AAP, BP and PJ are inventors of US patent number 9,782,411. CH received grants from Deutsche Forschungsgemeinschaft, Mitsubishi Cleansui, Wild Blueberry Association of North America, University of Surrey, Philips, National Processed Raspberry Council, Cranberry Institute and Rheacell, and personal fees from Bayer and Novo Nordisk, all outside of the submitted work.

### Data availability statement

All relevant preclinical and clinical data are provided within the paper and its supporting information files. Qualified researchers may request access to patient level data and related study documents including the clinical study report, blank case report form, statistical analysis plan, and dataset specifications. Patient level data will be anonymized, and study documents will be redacted to protect the privacy of trial participants. Further details on Sanofi’s data sharing criteria, eligible studies, and process for requesting access can be found at https://www.clinicalstudydatarequest.com.

### Supplementary materials

Supplemental Figure I. Body weight and fructosamine changes in diabetic rats.

Supplemental Figure II. Effect of SAR247799 (1 and 5 mg), sildenafil (50 mg), or placebo on systolic (A) and diastolic (B) blood pressure and pre-hyperemia brachial artery diameter (C) in type-2 diabetes patients.

## Notes

### Clinical Trial

NCT03462017

## REFERENCES

1. Rajendran, P. et al. The vascular endothelium and human diseases. Int J Biol Sci. 9, 1057–1069 (2013).

2. Taqueti, V. R. & Di Carli, M. F. Coronary microvascular disease: Pathogenic mechanisms and therapeutic options: JACC State-of-the-Art Review. J Am Coll Cardiol. 72, 2625–2641 (2018).

3. Marzilli, M. et al. Obstructive coronary atherosclerosis and ischemic heart disease: an elusive link! J Am Coll Cardiol. 60, 951–95 (2012).

4. van Sloten, T. T. et al. Endothelial dysfunction plays a key role in increasing cardiovascular risk in type 2 diabetes: the Hoorn study. Hypertension. 64, 1299–305 (2014).

5. Thuy, A. V., Reimann, C. M., Hemdan, N. Y. & Gräler, M. H. Sphingosine 1-Phosphate in blood: Function, metabolism, and fate. Cell Physiol Biochem. 34, 158–171 (2014)

6. Okajima, F. Plasma lipoproteins behave as carriers of extracellular sphingosine 1-phosphate: is this an atherogenic mediator or an anti-atherogenic mediator? Biochim Biophys Acta. 1582, 132–137 (2002).

7. Frej, C. et al. A Shift in ApoM/S1P between HDL-particles in women with type 1 diabetes mellitus is associated with impaired anti-inflammatory effects of the ApoM/S1P complex. Arterioscler Thromb Vasc Biol. 37, 1194–1205 (2017).

8. Vaisar, T. et al. Type 2 diabetes is associated with loss of HDL endothelium protective functions. PLoS One. doi:10.1371/journal.pone.0192616. eCollection (2018).

9. Brinck, J. W. et al. Diabetes mellitus is associated with reduced high-density lipoprotein sphingosine-1-phosphate content and impaired high-density lipoprotein cardiac cell protection. Arterioscler Thromb Vasc Biol. 36, 817–824 (2016).

10. Sattler, K. J. E. et al. Sphingosine 1-phosphate levels in plasma and HDL are altered in coronary artery disease. Basic Res Cardiol. 105, 821–832 (2010).

11. Argraves, K. et al. S1P, dihydro-S1P and C24:1-ceramide levels in the HDL-containing fraction of serum inversely correlate with occurrence of ischemic heart disease. Lipids Health Dis. doi:10.1186/1476-511X-10-70 (2011).

12. Knapp, M. et al. Plasma sphingosine-1-phosphate concentration is reduced in patients with myocardial infarction. Med Sci Monit. 15, CR490–493 (2009).

13. Knapp, M., Lisowska, A., Zabielski, P., Musiał, W. & Baranowski, M. Sustained decrease in plasma sphingosine-1-phosphate concentration and its accumulation in blood cells in acute myocardial infarction. Prostaglandins Other Lipid Mediat. 106, 53–61 (2013).

14. Denimal, D. et al. Impairment of the ability of HDL from patients with metabolic syndrome but without diabetes mellitus to activate eNOS: Correction by S1P enrichment. Arterioscler Thromb Vasc Biol. 37, 804–811 (2017).

15. Coldewey, S. M. et al. Elevation of serum sphingosine-1-phosphate attenuates impaired cardiac function in experimental sepsis. Sci Rep. 6, 27594. doi:10.1038/srep27594 (2016).

16. Winkler, M. S. et al. Loss of sphingosine 1-phosphate (S1P) in septic shock is predominantly caused by decreased levels of high-density lipoproteins (HDL). J Intensive care. 7. doi:10.1186/s40560-019-0376-2 (2019).

17. Jing, X. D. et al. The relationship between the high-density lipoprotein (HDL)-associated sphingosine-1-phosphate (S1P) and coronary in-stent restenosis Clinica Chimica Acta. 446, 248–252 (2015).

18. Soltau, I. et al. Serum sphingosine-1-phosphate concentrations are inversely associated with atherosclerotic diseases in humans. PloS One. e0168302. doi:10.1371/journal.pone.0168302. eCollection (2016).

19. Kiziltunc, E. et al. The relationship between pre-infarction angina and serum sphingosine-1-phosphate levels. Acta Cardiol Sin. 30, 546–552 (2014).

20. Kızıltunç, E. et al. Serum sphingosine 1 phosphate levels in patients with and without coronary collateral circulation. Acta Cardiol Sin. 34, 379–385 (2018).

21. Rosenson, R. S. et al. Dysfunctional HDL and atherosclerotic cardiovascular disease. Nat Rev Cardiol. 13, 48–60 (2016).

22. Armitage, J., Holmes, M. V. & Preiss, D. Cholesteryl ester transfer protein inhibition for preventing cardiovascular events: JACC Review Topic of the Week. J Am Coll Cardiol. 73, 477–487 (2019).

23. Boden, W. E. et al. Niacin in patients with low HDL cholesterol levels receiving intensive statin therapy. N Engl J Med. 365, 2255–2267 (2011).

24. Galvani, S. & Hla, T. Quality versus quantity: Making HDL great again. Arterioscler Thromb Vasc Biol. 37, 1018–1019 (2017).

25. Egom, E. E., Mamas, M. A. & Soran, H. HDL quality or cholesterol cargo: what really matters – spotlight on sphingosine-1-phophate-rich HDL Current opinion on lipidology. 24, 351–356 (2013).

26. Kurano, M. & Yatomi, Y. Sphingosine-1-phosphate and atherosclerosis J Atheroscler Thromb. 25, 16–26 (2018).

27. Argraves, K. M. et al. High density lipoprotein-associated sphingosine-1 phosphate promotes endothelial barrier function. J Biol Chem. 283, 25074–81 (2008).

28. Potì, F., Simoni, M. & Nofer, J-R. Atheroprotective role of high-density lipoprotein (HDL)-associated sphingosine-1-phosphate (S1P). Cardiovascular Research. 103, 395–404 (2014).

29. Ruiz, M. et al. High-density lipoprotein-associated apolipoprotein M limits endothelial inflammation by delivering sphingosine-1-phosphate to the sphingosine-1-phosphate receptor 1. Arterioscler Thromb Vasc Biol. 37, 118–129 (2017).

30. Sattler, J. K. E. Defects of High-density lipoproteins in coronary artery disease caused by low sphingosine-1-phosphate content. Correction by sphingosine-1-phosphate loading. J Am Coll Cardiol. 66, 1470–1485 (2015).

31. Christensen, P. M. et al. Impaired endothelial barrier function in apolipoprotein M–deficient mice is dependent on sphingosine-1-phosphate receptor 1. FASEB J. 30, 2351–2359 (2016).

32. Sanchez, T. Sphingosine-1-phosphate signaling in endothelial disorders. Curr Atheroscler Rep. 18, 31. doi:10.1007/s11883-016-0586-1 (2016).

33. Kappos, L. et al. FREEDOMS Study Group. A placebo-controlled trial of oral fingolimod in relapsing multiple sclerosis. N Engl J Med. 362, 387–401 (2010).

34. Kappos, L. et al. Siponimod versus placebo in secondary progressive multiple sclerosis (EXPAND): a double-blind, randomised, phase 3 study. Lancet. 391, 1263–1273 (2018).

35. Hoch, M., D’Ambrosio, D., Wilbraham, D., Brossard, P. & Dingemanse, J. Clinical pharmacology of ponesimod, a selective S1P₁ receptor modulator, after uptitration to supratherapeutic doses in healthy subjects. Eur J Pharm Sci. 63, 147–153 (2014).

36. Jain, N. & Bhatti, T. Fingolimod-associated macular edema: incidence, detection and management. Neurology. 78, 672–680 (2012).

37. Salvadori, M. et al. FTY720 versus MMF with cyclosporine in de novo renal transplantation: a 1-year, randomized controlled trial in Europe and Australasia, Am JTransplant. 6, 2912–2921 (2006).

38. Westhoff, T. H. et al. The impact of FTY720 (fingolimod) on vasodilatory function and arterial elasticity in renal transplant patients. Nephrol Dial Transplant. 22, 2354–2358 (2007).

39. Poirier, B., et al. (in press) A G-protein–biased S1P1 agonist, SAR247799, protects endothelial cells without affecting lymphocyte numbers. Science Signaling. (2020)

40. Thomas, N., Sweeney, K. & Somayaji, V. Meta-analysis of clinical dose-response in a large drug development portfolio. Stat Biopharmaceutical Res. 6, 302–317 (2014).

41. Clarkson, P. et al. Impaired vascular reactivity in insulin dependent diabetes mellitus is related to disease duration and low-density lipoprotein cholesterol levels. J Am Coll Cardiol. 28, 573–579 (1996).

42. Matsuzawa, Y., Kwon, T. G., Lennon, R. J., Lerman, L. O. & Lerman A. Prognostic value of flow-mediated vasodilation in brachial artery and fingertip artery for cardiovascular events: A systematic review and meta-analysis. J Am Heart Assoc. 4: doi:10.1161/JAHA.115.002270 (2015).

43. Aversa, A. et al. Chronic administration of Sildenafil improves markers of endothelial function in men with Type 2 diabetes. Diabet Med. 25, 37–44 (2008).

44. Balzer J. et al. Sustained benefits in vascular function through flavanol-containing cocoa in medicated diabetic patients. A double-masked, randomized, controlled trial. J. Am. Coll. Cardiol. 51, 2141–2149 (2008).

45. Desouza, C., Parulkar, A., Lumpkin, D., Akers, D. & Fonseca, V. Acute and prolonged effects of sildenafil on brachial artery flow-mediated dilatation in type 2 diabetes. Diabetes Care. 25, 1336–1339 (2002).

46. Deyoung, L., Chung, E., Kovac, J. R., Romano, W. & Brock, G. B. Daily use of sildenafil improves endothelial function in men with type 2 diabetes. J. Androl. 33, 176–180 (2012).

47. Halcox, J.P. et al. The effect of sildenafil on human vascular function, platelet activation, and myocardial ischemia J Am Coll Cardiol. 40, 1232–1240 (2002).

48. Rosano, G. M. et al. Chronic treatment with tadalafil improves endothelial function in men with cardiovascular risk. Eur Urol. 47, 214–222 (2005).

49. Shahin, Y., Khan, J. A., Samuel, N. & Cheeler, I. Angiotensin converting enzyme inhibitors effect on endothelial dysfunction: A meta-analysis of randomised controlled trials. Atherosclerosis. 216, 7–16 (2011).

50. Zhang, L., Gong, D., Li, S. & Zhou, X. Meta-analysis of the effects of statin therapy on endothelial function in patients with diabetes mellitus. Atherosclerosis. 223, 78–85 (2012).

51. Li, S., Wu, Y., Yu, G., Xia, Q. & Xu, Y. Angiotensin II receptor blockers improve peripheral endothelial function: a meta-analysis of randomized controlled trials. PloS one. doi:10.1371/journal.pone.0090217 (2014).

52. Clark, J. B., Palmer, C. J. & Shaw, W. N. The diabetic Zucker fatty rat. Proc Soc Exp Biol Med. 173, 68‐75 (1983).

53. Etgen, G. J. & Oldham BA. Profiling of Zucker diabetic fatty rats in their progression to the overt diabetic state. Metabolism. 49, 684‐688 (2000).

54. Ratcliffe, B., Pawlak, R., Morales, F., Harrison, C. & Gurovich, A. N. Internal validation of an automated system for brachial and femoral flow mediated dilation. Clin Hypertens. 23: 17. doi.org/10.1186/s40885-017-0073-1 (2017).

55. Stirban, A. et al. Acute effects of sildenafil on flow mediated dilatation and cardiovascular autonomic nerve function in type 2 diabetic patients. Diabetes Metab Res Rev. 25, 136–143. (2009).

56. Halcox, J. P. et al. Endothelial function predicts progression of carotid intima-media thickness. Circulation. 119, 1005–1012 (2009).

57. Ceconi, C. et al. ACE inhibition with perindopril and endothelial function. Results of a substudy of the EUROPA study: PERTINENT. Cardiovasc Res. 73, 237–246 (2007).

58. Taddei, S. et al. Effect of calcium antagonist or beta blockade treatment on nitric oxide-dependent vasodilation and oxidative stress in essential hypertensive patients. J Hypertens. 19, 1379–1386 (2001).

59. Rubinshtein, R. et al. Assessment of endothelial function by non-invasive peripheral arterial tonometry predicts late cardiovascular adverse events. Eur Heart J. 31, 1142–1148 (2010).

60. Barter, P. J. et al. Effects of torcetrapib in patients at high risk for coronary events. N Engl J Med. 357, 2109–2122 (2007).

61. Simic, B. et al. Torcetrapib impairs endothelial function in hypertension. Eur Heart J. 33, 1615–1624 (2012).

62. Lüscher, T. F. et al. Vascular effects and safety of dalcetrapib in patients with or at risk of coronary heart disease: the dal-VESSEL randomized clinical trial. Eur Heart J. 33, 857–865 (2012).

63. Schwartz, G. G. et al. Effects of dalcetrapib in patients with a recent acute coronary syndrome. N Engl J Med. 367, 2089–2099 (2012).

64. Charakida, M., Masi, S., Lüscher, T. F., Kastelein, J. J. & Deanfield, J. E. Assessment of atherosclerosis: the role of flow-mediated dilatation. Eur Heart J. 31, 2854–2861 (2010).

65. Igarashi, J. & Michel, T. Sphingosine-1-phosphate and modulation of vascular tone. Cardiovasc Res. 82, 212–212 (2009).

